# Novel Multiple Sclerosis Agents-Induced Cardiotoxicity

**DOI:** 10.1101/2021.12.11.21267656

**Authors:** Zaki Al-Yafeai, Hamzah Abduljabbar, Alexander Carvajal-González, Muhammed Arvas, Shaun Patel, Neev Patel

**Author notes:** <u>Corresponding Author:</u> Zaki Al-Yafeai MD, PhD, Internal Medicine Department.

## Abstract

**Background:** Emerging novel therapeutics have been developed to hamper the progression of multiple sclerosis. However, the adverse events related to these new agents remain largely unknown. Therefore, we sought to investigate the cardiovascular complications of these drugs.

**Methods:** Utilizing data from the U.S. Food and Drug Administration Adverse Events Reporting System, we comprehensively evaluated the cardiovascular complications of the newly FDA approved anti-multiple sclerosis agents. Disproportionality signal analysis was conducted by measuring reporting odds ratio (ROR) with 95% confidence interval of all the cardiovascular adverse events adverse events since approval till 2021.

**Results:** After vetting the newly approved agents for multiple sclerosis, CD20 and CD25 inhibitors and sphingosine-1-phosphate receptors agonists were the latest approved medications for multiple sclerosis since 2015. Two CD20 (ocrelizumab, ofatumumab) and one CD25 inhibitors (daclizumab) were significantly associated with multiple cardiovascular adverse events. Among all the cardiotoxic events; coronary artery disease, cardiac failure and atrial fibrillation were the most predominant among CD20 or CD25 blockers. Interestingly, Sphingosine-1-phosphate receptors agonists showed much fewer reported cardiac adverse events. However, fingolimod and siponimod were associated with significant bradycardia.

**Conclusions:** Our data revealed the new agents prescribed for multiple sclerosis have cardiotoxic effects, including not only the known adverse effects observed effects for S1P receptor modulators but also undefined cardiovascular complications associated with CD20 and CD25 inhibitors. These findings potentially instigate further studies to personalize prescribing these agents for multiple sclerosis based on patient’s cardiovascular profile.

## Introduction

Multiple sclerosis (MS) is known to be the most common neurologic demyelinating disease in young adults that it is characteristically a relapsing inflammatory demyelinating disease of the white matter. MS is rather complex inflammatory disorder that involves multiple immune cell lines, including macrophages, B cells, CD4+, CD8+, and T helper cells (Th 1, 2, 9, 17, 22)[1]. Initially a large body of literature supported the role of T cells in MS evident by earlier treatments for MS like Interferon B and Glatiramer acetate that act on these cells which were effective in decreasing relapses but not for the progression of the disease[2].However, the recently approved Sphingosine Phosphate 1 receptor (SPR1) modulator drugs have shown to be effective in decreasing the progression of the disease via limiting T-cell migration to tissues, and as a consequence, decreasing the immune response[3]. SPR1 are expressed in lymphocytes T and are part of the signaling process that promotes egress of lymphocytes from lymph nodes into the circulation[4]. Additionally, targeting CD25 on T cells using daclizumab has been proven to be effective for reducing MS relapses, however, due reported cases of autoimmune encephalitis the drug has been withdrawn[5].

Interestingly, over the last decade B cells have been consistently shown to drive a large portion of MS pathology[2, 6]. B cells harbor several antigens that are expressed at certain points of B cell evolution and play distinct role in its function and differentiation. Among these antigens, B cells express CD20 at most stages of development and differentiation. Due its presence in most B cell lineage, CD20 has been an attractive target for immunotherapy against B-cell lymphoma and leukemias[6]. Targeting CD20 either leads to accumulation of CD20 aggregates resulting in complement-mediated cytotoxicity or activation of apoptosis pathways. Evidently, CD20 antagonists limit MS relapses by depleting B cells, thus modulate the immune-inflammatory process of MS[7]. Several CD20 inhibitors have been approved by the FDA till now including ocrelizumab, ofatumumab, and rituximab[7, 8].

While rituximab cardiovascular adverse events have been reported, much less is known about the cardiotoxic effects of the other CD20 inhibitors, T-cell targeting therapy or S1P inhibitors. For example, the two large clinical trials (OPERA I and II) have established ocrelizumab as an effective therapy for multiple sclerosis. While multiple adverse events have been reported in these studies, cardiovascular complications were absent[9, 10]. Therefore, we sought to investigate the newly approved CD20, CD25 and S1P targeting agents and their cardiovascular complications.

## Methods

For this study we used the data available in the U.S Food and Drug Administration Adverse Reporting System (FAERS). This public pharmacovigilance monitoring database contains adverse reports events, medication error reports, and product quality complaints reported by health care professionals, manufacturers, and consumers worldwide since 1968; providing case identification numbers, name of the medication, adverse reaction, severity of the reaction, manufacturer of the pharmaceutical, concomitant pharmaceuticals and other related information. Since this is an anonymous database there was no need for ethical committee approval for this study.

In the present study we comprehensively evaluated the cardiovascular complications of the newly FDA approved anti-multiple sclerosis agents, selective sphingosine-1-phosphate receptor modulator (fingolimod, siponimod, ozanimod, ponesimod), humanized anti-CD20 monoclonal antibody (ocreluzimab, ofatumumab) and humanized anti CD-25 monoclonal antibody (Daclizumab). We looked for cardiovascular adverse events (AEs) using standardized medical terms according to the Medical Dictionary for Regulatory Activities, the list of terms included “atrial fibrillation”, “congestive cardiac failure, cardiac failure congestive”, “ventricular dysfunction”, “systolic dysfunction”; “diastolic dysfunction”, “cardiac disfunction”, “cardiac failure”, “cardiomyopathy”, “myocardial infarction”, “angina pectoris”, “coronary artery disease”, “coronary artery occlusion”, “angina unstable”, silent myocardial infarction, “acute coronary syndrome” and were organized in these main groups.

The association between the use of SPR1 modulators and humanized monoclonal antibodies with cardiovascular adverse events was assessed by disproportionality signal detection analysis using the reporting odds ratio (ROR). ROR is a measure of the magnitude of association between an exposure to a given pharmaceutical agent and the odds of patients experiencing a specific adverse event, compared to the odds of the same event occurring with all other pharmaceutical agents in the database. ROR was considered significant when the lower limit of the 95% confidence interval (CI) was >1.0.

## Results

The total number of AEs from all drugs in this study was 29,662. Amongst these, 25,014 were from the newly approved humanized monoclonal antibodies (daclizumab, ocrelizumab and ofatumumab) [Table 1 and Supplemental Tables 1-3]. These include 4,010 due to daclizumab, 17,018 due to ocrelizumab, and 3,986 were due to ofatumumab.

**Table 1.**
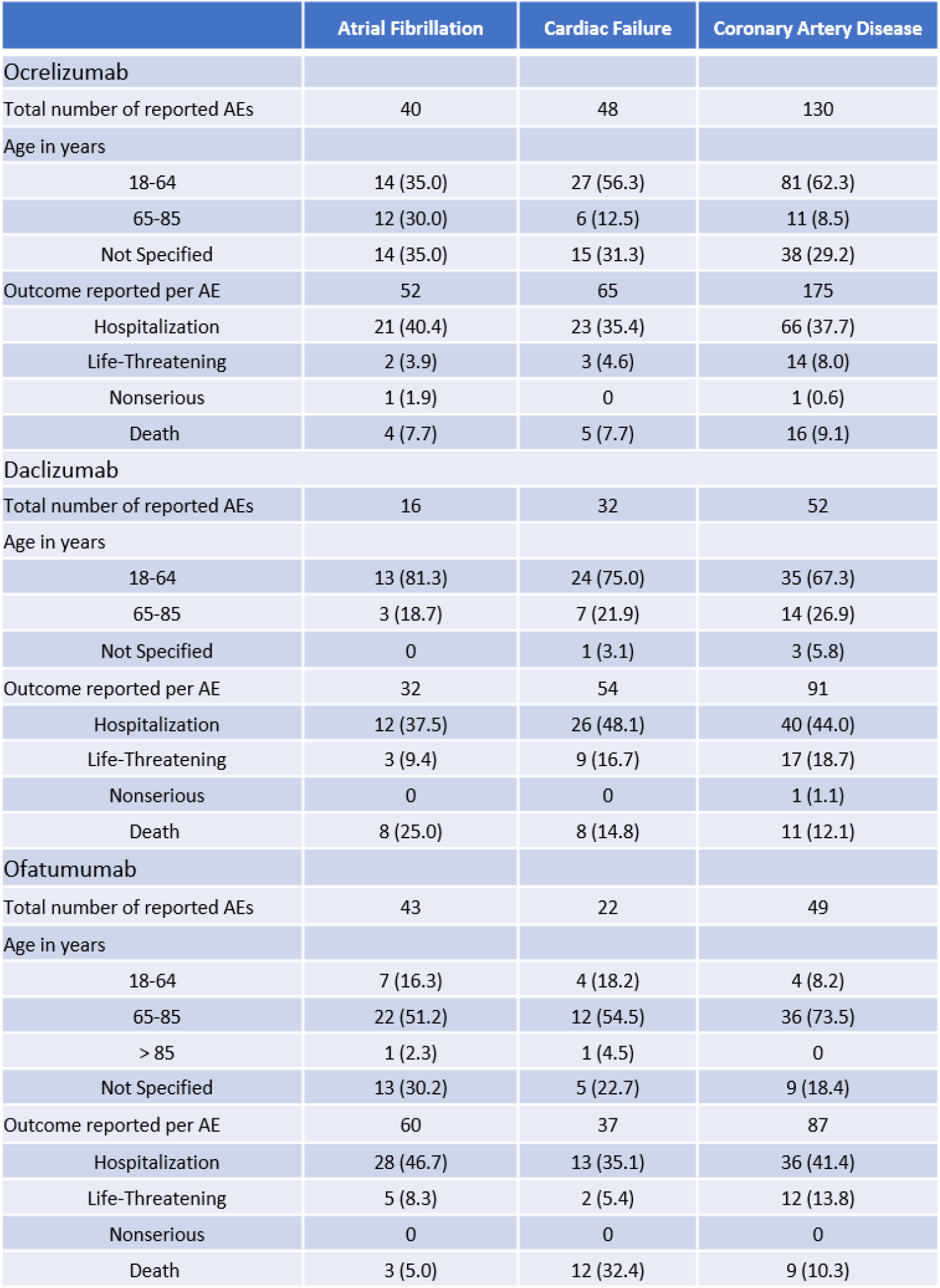
Values are n or n (%) unless otherwise indicated.

Amongst these reported AEs for daclizumab, ocrelizumab and ofatumumab, there were a total of 432 cardiac related AEs [Table 1]. A little over 50% of these cardiac related AEs were due to coronary artery disease. Ocrelizumab reported on the highest number of related CAD, but in relation to its total reported AEs had the lowest prevalence of 0.8% for CAD in comparison to daclizumab (1.5%) and ofatumumab (1.2%) [Table 2]. Heart failure was reported around 102 reported while 99 cases reported for atrial fibrillation associated with these three drugs [Table 1]. Ocrelizumab again had the highest reported cases for heart failure with a total of 48 reported AEs. For atrial fibrillation, the highest reported cases and the most prevalent occurred with ofatumumab, which had a total of 43 reported AEs.

**Table 2.** Values are n or n (%) unless otherwise indicated

| Total | Daclizumab only | Daclizumab +others | Daclizumab (total) | Ocrelizumab only | Ocrelizumab+ others | Ocrelizumab (total) | Ofatumumab only | Ofatumumab + others | Ofatumumab (total) |
| --- | --- | --- | --- | --- | --- | --- | --- | --- | --- |
| All AEs | 4010 |  |  | 17018 |  |  | 3986 |  |  |
| HF | 6 | 28 | 34 (0.85%) | 27 | 16 | 43 (0.3%) | 5 | 21 | 26 (0.65%) |
| AF | 1 | 15 | 16 (0.4%) | 27 | 8 | 35 (0.25%) | 7 | 34 | 41 (1%) |
| CAD | 10 | 49 | 59 (1.5%) | 89 | 16 | 105 (0.8%) | 14 | 34 | 48 (1.2%) |

Values are n or n (%) unless otherwise indicated

Tables 2 and Supplemental Tables 1-3. further characterize the cardiac related AEs that were associated with ocrelizumab, daclizumab, and ofatumumab. Ofatumumab and daclizumab showed male predominance (66% and 61%.4 respectively). Mortality rate was comparable among all the three medications. More than 90% of the cases were serious.

Next, we conducted disproportionality signal analysis for daclizumab, ocrelizumab and ofatumumab versus all other drugs in FAERS for CAD, cardiac failure and atrial fibrillation [Table 3]. Daclizumab reported the highest ROR for cardiac failure (2.9 with 95% CI: 2.07-4.07) with ofatumumab comes next (2.23, 95% CI:1.5-3.28). CAD showed the same pattern; daclizumab (ROR:3.8 with 95% CI: 2.96-4.96), ofatumumab (ROR: 3.1 with 95% CI: 2.33-4.12), and then ocrelizumab (ROR: 1.58 with 95% CI: 1.3-1.9). Interestingly, atrial fibrillation was only significantly elevated with ofatumumab is (ROR:3.06 with 95% CI: 2.25-4.17), however, ocrelizumab showed significantly low ROR (ROR: 0.5, 95% CI: 0.39-0.78). Since S1P receptor modulators showed negligible cardiac adverse events, disproportionality signal analysis was not done for this class of medications.

**Table 3.** Table 3 showing the reporting odds ratio (ROR) of daclizumab, ocrelizumab or ofatumumab vs all drugs in U.S. Food and Drug Administration Adverse Events Reporting System database regarding CAD, AF and cardiac failure. CAD: coronary artery disease. AF: atrial fibrillation.

| Drug name | Daclizumab vs Full Database | Ocrelizumab vs Full Database | Ofatumumab vs Full Database |
| --- | --- | --- | --- |
| CAD | 1.49 (0.62-1.06)<br>P = 0.1362 | 0.3975 (0.33-0.48)<br>P < 0.0001 | 0.5156 (0.36- 0.72) P = 0.0001 |
| Cardiac failure | 0.6399 (0.45 - 0.90)<br>P = 0.0103 | 0.1843 (0.1375 - 0.2472) P < 0.0001 | 0.3920 (0.25 - 0.60)<br>P < 0.0001 |
| AF | 0.3446 (0.21 - 0.56) P < 0.0001 | 0.1612(0.11 - 0.22) P < 0.0001 | 0.7322(0.52- 1.03)<br>P = 0.0737 |

Among the S1P receptor modulators (ponesimod, ozanimod, siponimod and fingolimod), there were a total of 4,648 reported AEs. [Supplemental Table 4]. Fingolimod showed the highest number of adverse events followed by siponimod. There was a total of 31 cardiac related events including atrial fibrillation, heart failure and coronary artery disease associated with siponimod while fingolimod had 927 cardiac related events. Siponimod and fingolimod showed a significant increase in bradycardia with ROR of 4.66 and 8.96, respectively [Table 4]. In addition, fingolimod also showed a significant association with AV block which was not observed for any of the other S1P receptor modulators.

**Table 4.** Table 4 showing the reporting odds ratio (ROR) of fingolimod, or siponimod vs all drugs in U.S. Food and Drug Administration Adverse Events Reporting System database regarding bradycardia and AV block. AV: atrioventricular.

| Drug name | Fingolimod vs others | Siponimod vs others |
| --- | --- | --- |
| Bradycardia | 8.9646 (8.6-9.4)<br>P<0.0001 | 4.66(3.7-5.9)<br>P<0.0001 |
| AV block | 2.9(2.7-3.2)<br>P<0.0001 | 1.45 (0.9-2.2)<br>P=0.08 |

## Discussion

The reported adverse events of the novel multiple sclerosis agents span multiple systems. While the cardiac related adverse events of CD20/CD25 inhibitors ranges from 3.45-5.8%, few major cardiac conditions were remarkably high. Specifically, our results showed that the risk of atrial fibrillation, cardiac failure and coronary artery disease were significantly elevated in patients taking daclizumab, ocrelizumab, ofatumumab for multiple sclerosis. Interestingly, coronary artery disease showed the highest ROR compared to the other major adverse events. However, when compared to other CD20 inhibitors (ibritumomab, rituximab, obinutuzumab), daclizumab, ocrelizumab, ofatumumab were associated with significantly lower ROR regarding atrial fibrillation, cardiac failure and coronary artery disease.

Over the last two decades, the role of immune cell depletion via CD20/CD25 inhibitors received considerable attention in the field of neuroimmunology. Rituximab probably was the first CD20 inhibitors used for multiple sclerosis, however, its cardiovascular complications are well studied. Almost 10 years later, two major clinical trials (OPERA I and OPERA II) investigated ocrelizumab in relapsing remitting multiple sclerosis and elegantly revealed the therapeutic potential of ocrelizumab. Subsequent clinical trials placed ofatumumab in therapeutic array for multiple sclerosis. Unfortunately, Daclizumab approval for multiple sclerosis vanished quickly after serious adverse forced the company to withdraw the drug from the market.

While clinical studies demonstrated the cardiotoxicity of rituximab (e.g. atrial fibrillation)[11, 12], much less is known about the new immunomodulators associated cardiotoxicity. Clinical trials of ocrelizumab for multiple sclerosis showed no serious adverse events[9, 10, 13]. However, trials for rheumatoid arthritis were actually terminated after the STAGE study showed one patient died due to myocardial infarction after receiving 500 mg (higher dose when compared to multiple sclerosis dose)[14, 15]. Early clinical trials on daclizumab reported rare cases of allergic myonecrosis of the myocardium and pleural effusions[16]. Two cases of congestive heart failure have been reported in patients taking daclizumab for adult T-cell leukemia[5]. One case of atrial fibrillation has been linked to Ofatumumab in a trial comparing ofatumumab versus Ibrutinib[17]. Even rarer (<1%), pericarditis, heart failure and myocardial infarction cases were reported to be associated with ofatumumab use[18].

Fingolimod, the first SPR modulator introduced into the market, is well known for its cardiovascular side effects, mainly bradycardia and AV block. The FREEDOMS and TRANSFORMS trials reported respectively 14 (3%) and 9 (2.1%) cases of bradycardia in patients on the 1.25 mg dose[19, 20], which were corroborated in our study with a significantly higher ROR compared to other SPR1 modulator. These side effects are thought to be mediated by the activation of S1P1 and S1P3 receptors in cardiomyocytes[21]. Interestingly, siponimod, a newer SPR1 modulator that binds more selectively to S1P1 and S1P5, was expected to have lower risk of bradycardia and AV block, however, our study shows that siponimod also has a significantly high ROR for bradycardia compared to the other SPR agents. While ponesimod and ozanimod did not show high incidence of bradycardia or AV block suggesting that these agents might be more specific.

Despite of the novelty of our findings, certain limitations to this study are appreciated. For example, patient’s comorbidities and other risk factors are unknown. Additionally, cardiovascular history and baseline cardiac functioning are not taken into account when performing these studies. Besides patient’s risk stratification, the molecular mechanisms underlying immunomodulators-associated cardiotoxicity remain unknown.

The importance of this study comes from the wide use of these medications for multiple sclerosis management. While significant studies addressed few cardiac adverse events of the novel MS therapy as mentioned earlier, comprehensive cardiotoxicity studies are wanting. The present study corroborates with the findings that novel MS therapy does indeed have cardiotoxicity risk. As the use of these new agents increases for MS treatment, the need for cardiovascular risk stratification is much needed now than ever. Since the risk of cardiac failure, coronary artery disease, atrial fibrillation, and bradycardia is enhanced in patients using CD20/CD25 inhibitors and S1P modulators, we suggest clinician to obtain baseline electrocardiography and echocardiogram when initiating these medications. Additionally, patients should be educated about the cardiac risk when taking these medications. Physician are recommended to monitor the sign and symptoms of cardiac failure and coronary artery disease during the treatment period.

## Data Availability

All of the data in the current study is available in the manuscript

## Acknowledgment

None.

