## Supplemental data for "Novel Multiple Sclerosis Agents-Induced Cardiotoxicity"

### Slide 1
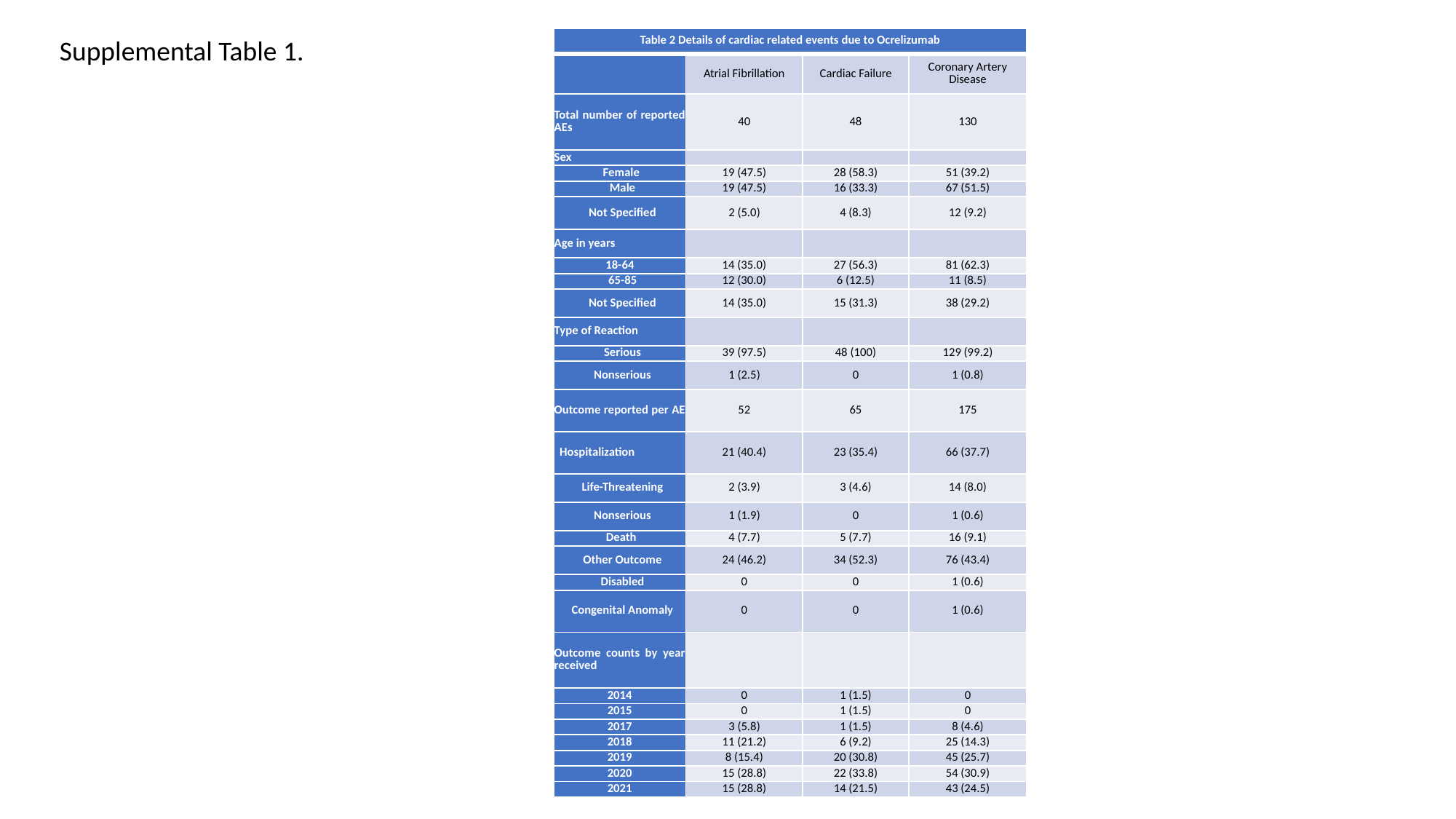

Supplemental Table 1.
| Table 2 Details of cardiac related events due to Ocrelizumab | | | |
| --- | --- | --- | --- |
| | Atrial Fibrillation | Cardiac Failure | Coronary Artery Disease |
| Total number of reported AEs | 40 | 48 | 130 |
| Sex | | | |
| Female | 19 (47.5) | 28 (58.3) | 51 (39.2) |
| Male | 19 (47.5) | 16 (33.3) | 67 (51.5) |
| Not Specified | 2 (5.0) | 4 (8.3) | 12 (9.2) |
| Age in years | | | |
| 18-64 | 14 (35.0) | 27 (56.3) | 81 (62.3) |
| 65-85 | 12 (30.0) | 6 (12.5) | 11 (8.5) |
| Not Specified | 14 (35.0) | 15 (31.3) | 38 (29.2) |
| Type of Reaction | | | |
| Serious | 39 (97.5) | 48 (100) | 129 (99.2) |
| Nonserious | 1 (2.5) | 0 | 1 (0.8) |
| Outcome reported per AE | 52 | 65 | 175 |
| Hospitalization | 21 (40.4) | 23 (35.4) | 66 (37.7) |
| Life-Threatening | 2 (3.9) | 3 (4.6) | 14 (8.0) |
| Nonserious | 1 (1.9) | 0 | 1 (0.6) |
| Death | 4 (7.7) | 5 (7.7) | 16 (9.1) |
| Other Outcome | 24 (46.2) | 34 (52.3) | 76 (43.4) |
| Disabled | 0 | 0 | 1 (0.6) |
| Congenital Anomaly | 0 | 0 | 1 (0.6) |
| Outcome counts by year received | | | |
| 2014 | 0 | 1 (1.5) | 0 |
| 2015 | 0 | 1 (1.5) | 0 |
| 2017 | 3 (5.8) | 1 (1.5) | 8 (4.6) |
| 2018 | 11 (21.2) | 6 (9.2) | 25 (14.3) |
| 2019 | 8 (15.4) | 20 (30.8) | 45 (25.7) |
| 2020 | 15 (28.8) | 22 (33.8) | 54 (30.9) |
| 2021 | 15 (28.8) | 14 (21.5) | 43 (24.5) |

### Slide 2
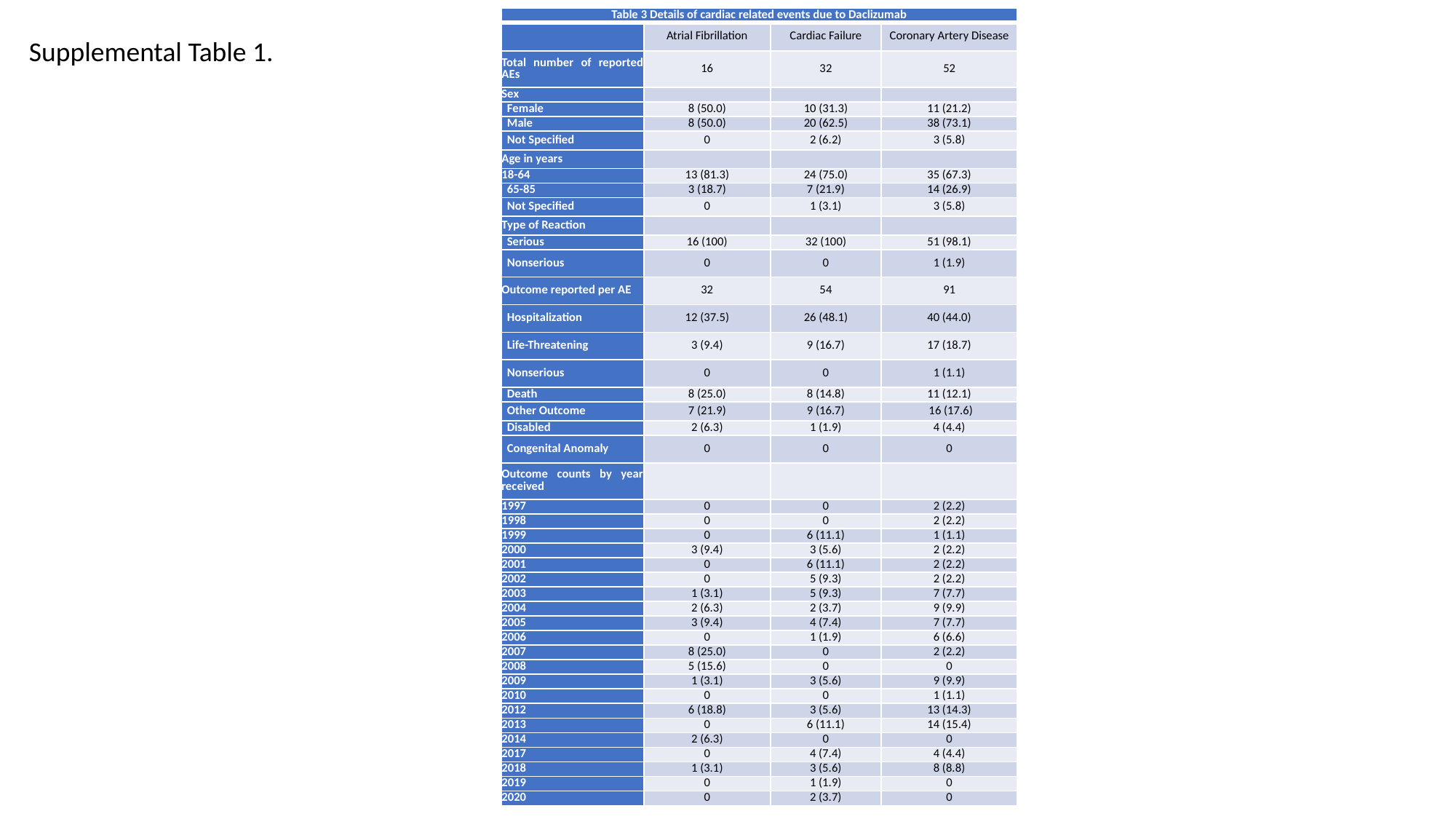

| Table 3 Details of cardiac related events due to Daclizumab | | | |
| --- | --- | --- | --- |
| | Atrial Fibrillation | Cardiac Failure | Coronary Artery Disease |
| Total number of reported AEs | 16 | 32 | 52 |
| Sex | | | |
| Female | 8 (50.0) | 10 (31.3) | 11 (21.2) |
| Male | 8 (50.0) | 20 (62.5) | 38 (73.1) |
| Not Specified | 0 | 2 (6.2) | 3 (5.8) |
| Age in years | | | |
| 18-64 | 13 (81.3) | 24 (75.0) | 35 (67.3) |
| 65-85 | 3 (18.7) | 7 (21.9) | 14 (26.9) |
| Not Specified | 0 | 1 (3.1) | 3 (5.8) |
| Type of Reaction | | | |
| Serious | 16 (100) | 32 (100) | 51 (98.1) |
| Nonserious | 0 | 0 | 1 (1.9) |
| Outcome reported per AE | 32 | 54 | 91 |
| Hospitalization | 12 (37.5) | 26 (48.1) | 40 (44.0) |
| Life-Threatening | 3 (9.4) | 9 (16.7) | 17 (18.7) |
| Nonserious | 0 | 0 | 1 (1.1) |
| Death | 8 (25.0) | 8 (14.8) | 11 (12.1) |
| Other Outcome | 7 (21.9) | 9 (16.7) | 16 (17.6) |
| Disabled | 2 (6.3) | 1 (1.9) | 4 (4.4) |
| Congenital Anomaly | 0 | 0 | 0 |
| Outcome counts by year received | | | |
| 1997 | 0 | 0 | 2 (2.2) |
| 1998 | 0 | 0 | 2 (2.2) |
| 1999 | 0 | 6 (11.1) | 1 (1.1) |
| 2000 | 3 (9.4) | 3 (5.6) | 2 (2.2) |
| 2001 | 0 | 6 (11.1) | 2 (2.2) |
| 2002 | 0 | 5 (9.3) | 2 (2.2) |
| 2003 | 1 (3.1) | 5 (9.3) | 7 (7.7) |
| 2004 | 2 (6.3) | 2 (3.7) | 9 (9.9) |
| 2005 | 3 (9.4) | 4 (7.4) | 7 (7.7) |
| 2006 | 0 | 1 (1.9) | 6 (6.6) |
| 2007 | 8 (25.0) | 0 | 2 (2.2) |
| 2008 | 5 (15.6) | 0 | 0 |
| 2009 | 1 (3.1) | 3 (5.6) | 9 (9.9) |
| 2010 | 0 | 0 | 1 (1.1) |
| 2012 | 6 (18.8) | 3 (5.6) | 13 (14.3) |
| 2013 | 0 | 6 (11.1) | 14 (15.4) |
| 2014 | 2 (6.3) | 0 | 0 |
| 2017 | 0 | 4 (7.4) | 4 (4.4) |
| 2018 | 1 (3.1) | 3 (5.6) | 8 (8.8) |
| 2019 | 0 | 1 (1.9) | 0 |
| 2020 | 0 | 2 (3.7) | 0 |
Supplemental Table 1.

### Slide 3
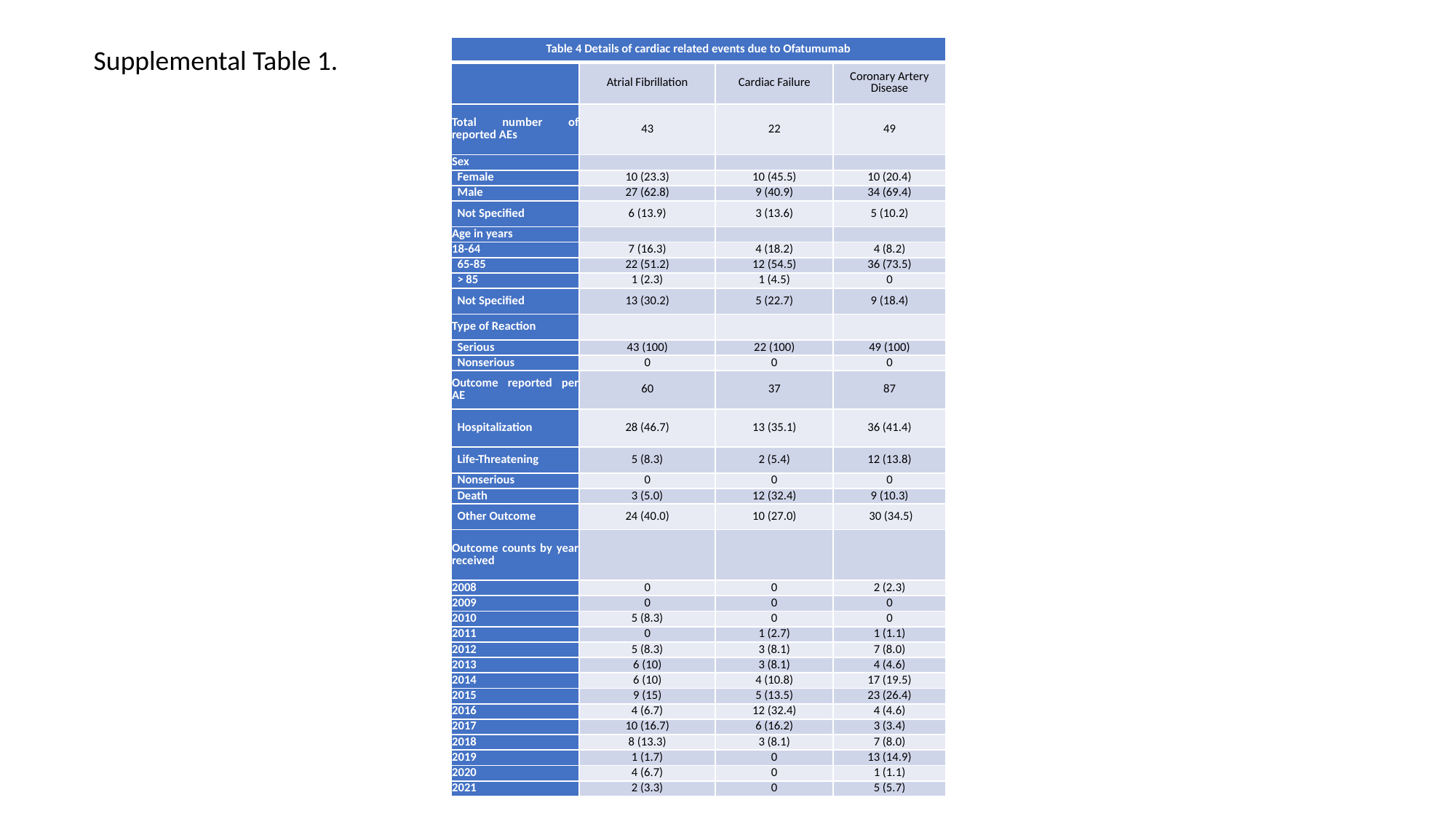

Supplemental Table 1.
| Table 4 Details of cardiac related events due to Ofatumumab | | | |
| --- | --- | --- | --- |
| | Atrial Fibrillation | Cardiac Failure | Coronary Artery Disease |
| Total number of reported AEs | 43 | 22 | 49 |
| Sex | | | |
| Female | 10 (23.3) | 10 (45.5) | 10 (20.4) |
| Male | 27 (62.8) | 9 (40.9) | 34 (69.4) |
| Not Specified | 6 (13.9) | 3 (13.6) | 5 (10.2) |
| Age in years | | | |
| 18-64 | 7 (16.3) | 4 (18.2) | 4 (8.2) |
| 65-85 | 22 (51.2) | 12 (54.5) | 36 (73.5) |
| > 85 | 1 (2.3) | 1 (4.5) | 0 |
| Not Specified | 13 (30.2) | 5 (22.7) | 9 (18.4) |
| Type of Reaction | | | |
| Serious | 43 (100) | 22 (100) | 49 (100) |
| Nonserious | 0 | 0 | 0 |
| Outcome reported per AE | 60 | 37 | 87 |
| Hospitalization | 28 (46.7) | 13 (35.1) | 36 (41.4) |
| Life-Threatening | 5 (8.3) | 2 (5.4) | 12 (13.8) |
| Nonserious | 0 | 0 | 0 |
| Death | 3 (5.0) | 12 (32.4) | 9 (10.3) |
| Other Outcome | 24 (40.0) | 10 (27.0) | 30 (34.5) |
| Outcome counts by year received | | | |
| 2008 | 0 | 0 | 2 (2.3) |
| 2009 | 0 | 0 | 0 |
| 2010 | 5 (8.3) | 0 | 0 |
| 2011 | 0 | 1 (2.7) | 1 (1.1) |
| 2012 | 5 (8.3) | 3 (8.1) | 7 (8.0) |
| 2013 | 6 (10) | 3 (8.1) | 4 (4.6) |
| 2014 | 6 (10) | 4 (10.8) | 17 (19.5) |
| 2015 | 9 (15) | 5 (13.5) | 23 (26.4) |
| 2016 | 4 (6.7) | 12 (32.4) | 4 (4.6) |
| 2017 | 10 (16.7) | 6 (16.2) | 3 (3.4) |
| 2018 | 8 (13.3) | 3 (8.1) | 7 (8.0) |
| 2019 | 1 (1.7) | 0 | 13 (14.9) |
| 2020 | 4 (6.7) | 0 | 1 (1.1) |
| 2021 | 2 (3.3) | 0 | 5 (5.7) |

### Slide 4
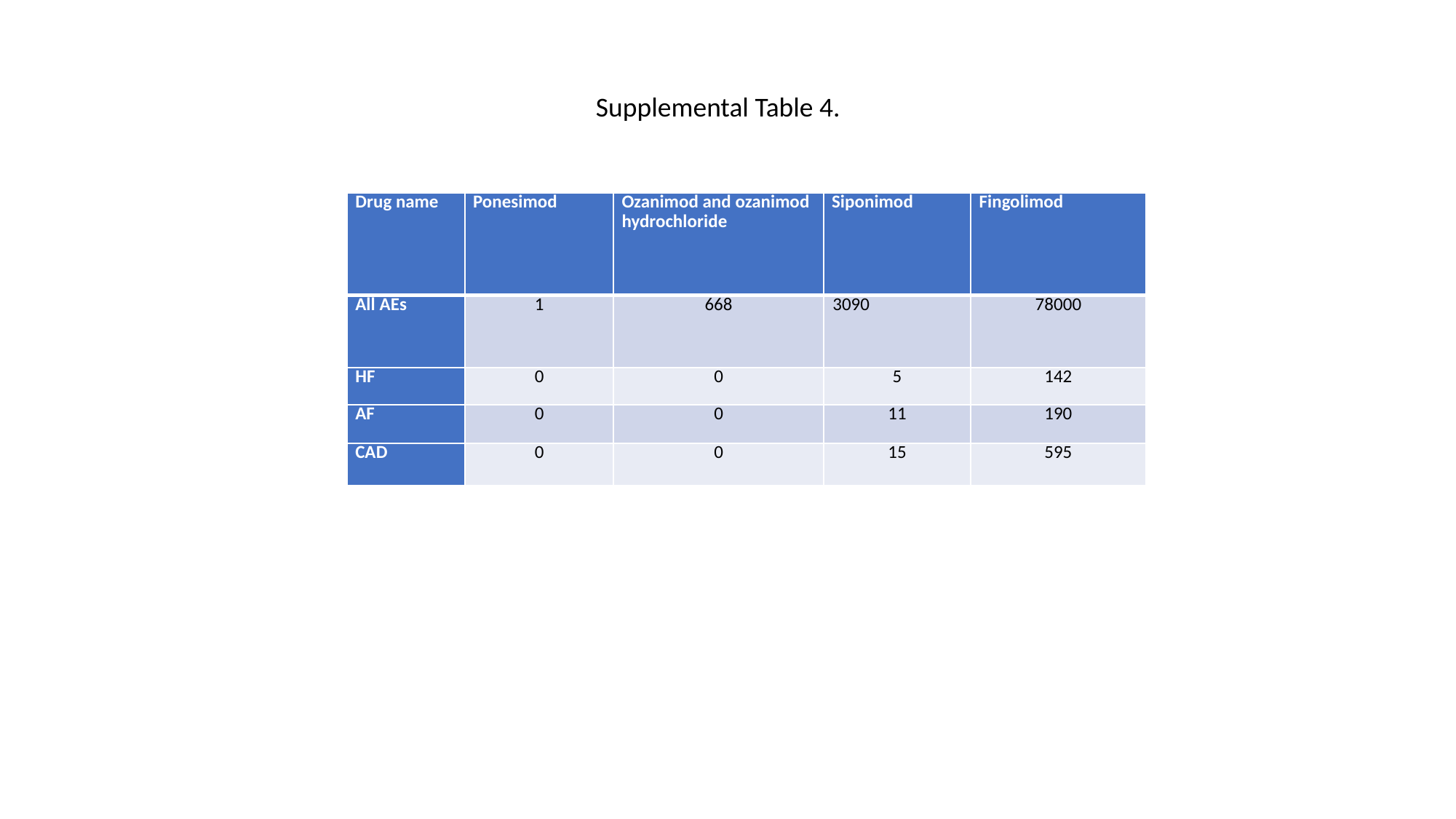

Supplemental Table 4.
| Drug name | Ponesimod | Ozanimod and ozanimod hydrochloride | Siponimod | Fingolimod |
| --- | --- | --- | --- | --- |
| All AEs | 1 | 668 | 3090 | 78000 |
| HF | 0 | 0 | 5 | 142 |
| AF | 0 | 0 | 11 | 190 |
| CAD | 0 | 0 | 15 | 595 |
